# The relatively young and rural population may limit the spread and severity of Covid-19 in Africa: a modelling study

**DOI:** 10.1101/2020.05.03.20089532

**Authors:** Binta Zahra Diop, Marième Ngom, Clémence Pougué Biyong, John N. Pougué Biyong

## Abstract

**Introduction:** A novel coronavirus disease 2019 (COVID-19) has spread to all regions of the world. There is great uncertainty regarding how countries characteristics will affect the spread of the epidemic; to date, there are few studies that attempt to predict the spread of the epidemic in African countries. In this paper, we investigate the role of demographic patterns, urbanization and co-morbidities on the possible trajectories of COVID-19 in Ghana, Kenya, and Senegal.

**Methods:** We use an augmented deterministic SIR model to predict the true spread of the disease, under the containment measures taken so far. We dis-aggregate the infected compartment into asymptomatic, mildly symptomatic, and severely symptomatic to match observed clinical development of COVID-19. We also account for age structures, urbanization, and co-morbidities (HIV, tuberculosis, anemia).

**Results:** In our baseline model, we project that the peak of active cases will occur in July, subject to the effectiveness of policy measures. When accounting for the urbanization, and factoring-in co-morbidities, the peak may occur between June 2^nd^ and June 17^th^ (Ghana), July 22^nd^ and August 29^th^ (Kenya), and finally May 28^th^ and June 15^th^ (Senegal). Successful containment policies could lead to lower rates of severe infections. While most cases will be mild, we project in the absence of policies further containing the spread, that between 0.78 and 1.03%, 0.61 and 1.22%, and 0.60 and 0.84% of individuals in Ghana, Kenya, and Senegal respectively may develop severe symptoms at the time of the peak of the epidemic.

**Conclusion:** Compared to Europe, Africa’s younger and rural population may modify the severity of the epidemic. The large youth population may lead to more infections but most of these infections will be asymptomatic or mild, and will probably go undetected. The higher prevalence of underlying conditions must be considered.

**Summary:** *What is known?:*

- While most COVID-19 studies focus on western and Asian countries, very few are concerned with the spread of the virus in African countries.
- Most African countries have relatively low urbanization rates, a young population and context-specific co-morbidities that are still to be explored in the spread of COVID-19.

*What are the new findings?:*

- In our baseline predictions 33 to 50% of the public will be actively infected at the peak of the epidemic and 1 in 36 (Ghana), 1 in 40 (Kenya) and 1 in 42 (Senegal) of these active cases may be severe.
- With rural areas, infection may be lowered to 65-73% (Ghana), 48-71% (Kenya) and 61-69% (Senegal) of the baseline infections.
- Comorbidities may however increase the ratio of severe infections among the active cases at the peak of the epidemic.

*What do the new findings imply?:*

- Rural areas and large youth population may limit the spread and severity of the epidemic and outweigh the negative impact of HIV, tuberculosis and anemia.

## INTRODUCTION

Since the first reported severe acute syndrome coronavirus 2 (SARS-CoV-2) infection in December 2019, the virus has spread to all continents.[1] There is still little evidence on the pattern of the spread in Africa. Although the African continent is made up of countries with different infrastructures, health policies, and characteristics in the face of this novel coronavirus disease 2019 (COVID-19); some characteristics such as a young population,[2] co-morbidities (tuberculosis, HIV, anemia[3,4]) and low urbanization rates transcend these differences and have been seldom considered in the large number of studies published to date. For example, the median age below 20,[5] and the low rates of urbanization, could potentially lead to a lower death toll of the epidemic in African countries than elsewhere.

However, having a young population implies that many infected individuals may not display symptoms and will risk infecting more people than would symptomatic individuals.[6] Additionally, the large number of informal settlements could accentuate this phenomenon. It is therefore urgent to develop a framework that could accurately predict the spread of the virus, accounting for the idiosyncrasies of the African context. A country-specific model will provide policy makers with a wide range of prediction scenarios, based on different actions they can take to address the pandemic. With the scarce resources at their disposal,[7,8] models like these will help target prevention strategies to individuals with co-morbidities who might suffer the most from the epidemic. Moreover, with containment policies that can grind economies to a halt,[9] understanding the trade-off between rural and urban spreads could lead to better informed decisions between the short term impacts of the epidemic, and the long-term looming shortage in the food-supply that could stem enforcing strict social distancing measures in rural areas.

This study contributes to the meager literature on the burden of the virus on African countries; it also adds to the use of differential equation models to predict the spread of epidemics. This paper focuses on three African countries that have received little attention: Ghana, Kenya, and Senegal. We chose Kenya to have a comparison point with another in-depth study by Brand et al.[10] Ghana and Senegal on the other hand have had transparent data sharing policies from the start of the epidemic; they made available publicly the number of positive cases, the number tests conducted and a clear outline of the containment measures. Ghana has an extensive testing policy, while Senegal has tested very few individuals comparatively^1^, we are thus able to see the difference in predictions for two countries that have adopted widely different testing strategies.

To project the trends of the epidemic, we augment the canonical *Susceptible - Infected - Recovered* by splitting the *infected* compartment into three groups: an infected without symptoms, an infected with mild symptoms, and finally, the infected with severe symptoms. With our projections accounting for policies implemented to date, we present different scenarios accounting for local policies, urbanization, and co-morbidities. Our strategy is relevant beyond the application of this paper; it could be used in Asian or European contexts as well, and is similar to work by Ferguson et al.[12] who discuss suppression and mitigation measures in the UK and the USA.

## METHODS

### Compartmental epidemiological model

Several models have been used to predict the spread of the virus. Read et al.[13] use a standard *Susceptible – Exposed – Infected – Recovered* (SEIR) model with an *exposed* compartment that comprises infected individuals who do not yet have symptoms and who are not infectious. Danon et al. [14] also use a SEIR model but split the *infected* compartment into two sub-compartments: mild symptoms and symptomatic. Finally, Arenas et al.[15] study use a model composed of *susceptible, exposed, asymptomatic infectious, infected, hospitalized to ICU, dead*, and *recovered* compartments; however, they assume that all asymptomatic infectious individuals cannot recover before they ever develop symptoms.

There is early evidence that a large number of individuals infected with COVID-19 will recover without ever developing symptoms and that asymptomatic individuals are contagious to varying degrees.[16–19] Based on these findings, our model assumes that individuals are contagious from the moment they get infected. We define a *Susceptible - Infected – Recovered* (SIR) model with vital dynamics (see figure 1 of the appendix). The known natural progression of the disease is (1) asymptomatic, (2) mild symptomatic, (3) moderate symptomatic, (4) severe, and (5) critical. There are benefits in understanding the heterogeneity among infected individuals namely, those carriers without symptoms (asymptomatic), carriers without symptoms (mild and moderately symptomatic), and severe cases who might seek medical attention (severe and critical). We therefore propose to divide the *infected* compartment into three sub-compartments: asymptomatic infectious, mildly (and moderate) symptomatic infectious, and severely (and critically) infected requiring medical attention.

We introduce some notations. *S* is the share of susceptible, i.e. individuals who are exposed to the virus but not immune. *I_as_*, *I_ms_*, and *I_ss_* are respectively the shares of asymptomatic, mildly symptomatic, and severely symptomatic individuals. *R* is the share of immune individuals. *D* is the share of deceased individuals (due to COVID-19 and other non-related causes). All numbers are expressed in terms of percentages of the total population. We note *I* = *I_as_* + *I_ms_* + *I_ss_* the total share of *infected* and *N* = *S* + *I* + *R* the share of individuals alive. We suppose that borders are closed. Moreover, all compartments experience natural vital dynamics via the birth rate *μ_birth_* and the death rate *μ_death_* from causes unrelated to the virus (e.g. long-term diseases, accidents). Daily epidemic transmission is described by equations (1)-(5):

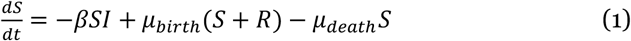

where *βSI* = *β_as_ SI_as_ +β_ms_SI_ms_* + *β_ss_SI_ss_*.

*β_as_* is the contact rate between asymptomatic *infected* individuals and *susceptible* ones. Because asymptomatic individuals are not aware of their infection, their rate of contact with susceptible individuals is the same as the rate of contact within the group of *susceptible* individuals. This contact rate will vary with containment measures that are enforced within each country. We define the asymptomatic effective reproduction number *R_t_* = *β_as_ * T_rec,as_* as the average number of secondary cases per asymptomatic case at time *t*.

*β_ms_* is the contact rate between mildly symptomatic individuals and susceptible ones. It is assumed to be lower than *β_as_* because symptomatic individuals tend to self-isolate, either because they are bedridden due to their symptoms or simply because they want to limit contacts with *susceptible* individuals.

*β_ss_* is the contact rate between severely symptomatic and *susceptible* individuals. Individuals who experience severe symptoms may seek medical care and get admitted as inpatients at a hospital. They might not get hospital care for various reasons (e.g. health facilities are overwhelmed). This rate accounts for contacts between hospitalized patients and healthcare workers, but can also be interpreted as contacts between severely symptomatic individuals and any care-giver (at home for instance, if the health services are overloaded). It also accounts for the contacts between hospitalized severely symptomatic individuals and *susceptible* individuals outside of their care-takers. It remains unclear how contacts other than healthcare workers affect the value of *β_ss_*.

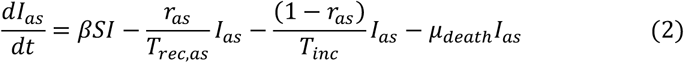

where *r_as_* is the probability of recovery without ever developing symptoms, *T_rec,as_* is the recovery time of an asymptomatic individual, and *T_inc_* is the incubation period during which an individual is infected and infectious, but does not have symptoms.

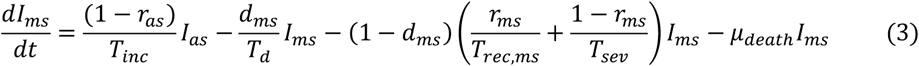

where *d_ms_* is the probability of dying from a fast deterioration, *T_d_* is the time elapsed between the appearance of first symptoms and the death of the individual, *r_ms_* is the probability to recover from mild symptoms, *T_rec,ms_* is the recovery time associated with *r_ms_* and *T_sev_* is the time for severe symptoms to develop. We deviate for the recovery rates of the mildly symptomatic compartment *r_ms_* by taking the weighted average of age-grouped fatality rates of COVID-19 found in Hubei, Hong Kong, and Macau [40]:

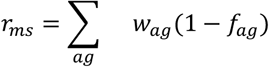

Where the sum is over the age groups *ag* ∈ {[0, 9], [10, 19],…, [70, 79], 80+}, *w_ag_* is the share of the population in age group *ag* and *f_ag_* is the fatality rate found in earlier studies for the population in age group [18].

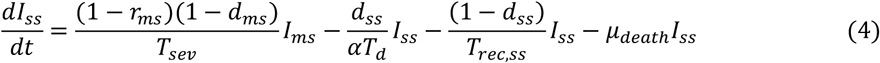

Where *d_ss_* is the probability of dying after progressively developing severe symptoms that require hospitalization, *T_rec,ss_* is the recovery time of the severely symptomatic. *αT_d_* is the time to death from the start of severe symptoms, for individuals who pass away from severe, progressively developing symptoms. Intuitively, if most severe cases are hospitalized, *α* should be higher than 1 as health professionals will slow down the evolution of the disease.

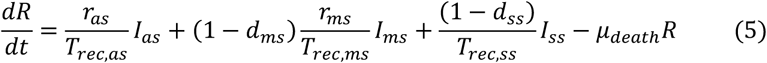

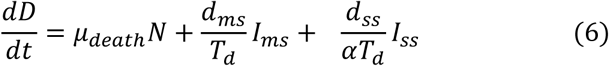

In our simulations, we include fatalities, but we do not include the outcomes in the results. We make this choice because of the high uncertainty around the capacity of the healthcare systems of each individual country to absorb the increased demand from severely ill patients. For instance, with the same predictions, a country that has a stock of ventilators of 1,000 will likely have less fatalities than a country with no ventilators. Because we do not have data on healthcare capacities, therefore we chose not to present these results. Our predicted number of fatalities are however subtracted from the number of susceptibles.

### Patient and Public Involvement

Our study does not involve the participation of patients or any members of the public. All data used for the purpose of this study are aggregated and publicly available.

## RESULTS

### Baseline Simulations

We use publicly available data from the European Center for Disease Control and Prevention, and from daily press releases made by the Senegalese ministry of health and social protection. [20,21] We also do checks using the Ghana Health Service and the Kenya ministry of health websites.

Whenever possible, we use values of parameters drawn from the literature to fit the model (see table 1).

Although there are reports that as many as 80% of active cases are asymptomatic,[22] these reports are based on cases that are still active and include pre-symptomatic individuals. We thus use 40% as the share of individuals infected with COVID-19 who recover without ever developing symptoms.[16–19]

Ghana, Kenya, and Senegal have extensive communication strategies including in local languages to ensure that communities are able to detect the symptoms of COVID-19 such as a cough and fever and would report any person with those symptoms. We there use the share of individuals who do not have a cough as a proxy for the rate of symptomatic infected individuals who can leave their home without being reported. Wang et al.[23] find that 59% of individuals who test positive for COVID-19 have a cough which implies *β_ms_* = 0.41*β_as_*. We set 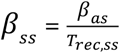 we presume that individuals are most at risk of infecting other susceptibles during their transport to the hospital. This ratio therefore mean that a severely ill patient would infect as many individuals over the course of their severely symptomatic phase as an asymptomatic individual would over the course of one day.

We chose South-Korea as a benchmark to validate *β_ms_*, *β_ss_*, and other parameters of our model because it is cited as an example for its extensive testing, tracking, and tracing of infections. We calibrate *R_t_* by allowing it to change at each new containment measure taken by South-Korean authorities until the number of identified cases reach a plateau. We solve an optimization problem constrained by (1)-(5) using the MATLAB optimization codes of D’Errico.[24] The values of *R_t_* obtained and other parameters are summarized in table 1. The number of infections computed with our model accurately approximates the positive cases in South-Korea (see figure 1).

For the fatality rates *d_ss_* and *d_ms_* in Ghana, Kenya and Senegal, we split the reported regional fatality rate^2^ 2.37% between *d_ss_* = 2% and *d_ms_=*0.37% as most deaths occurred for the severely symptomatic^3^.[25] However, in South Korea, we use the South Korean COVID-19 fatality rate — 1.07 % as of April 2, 2020 — and split it across *d_ms_*=0.03% and *d_ss_*=1.04%.

*I_ms_* was initialized with the number of cases tested positive on the first day of the epidemic in the country. *I_as_* was initialized with the number of cases *T_inc_* = 5 days after this same date. *I_ss_* was initialized at 0.

**Figure 1:**
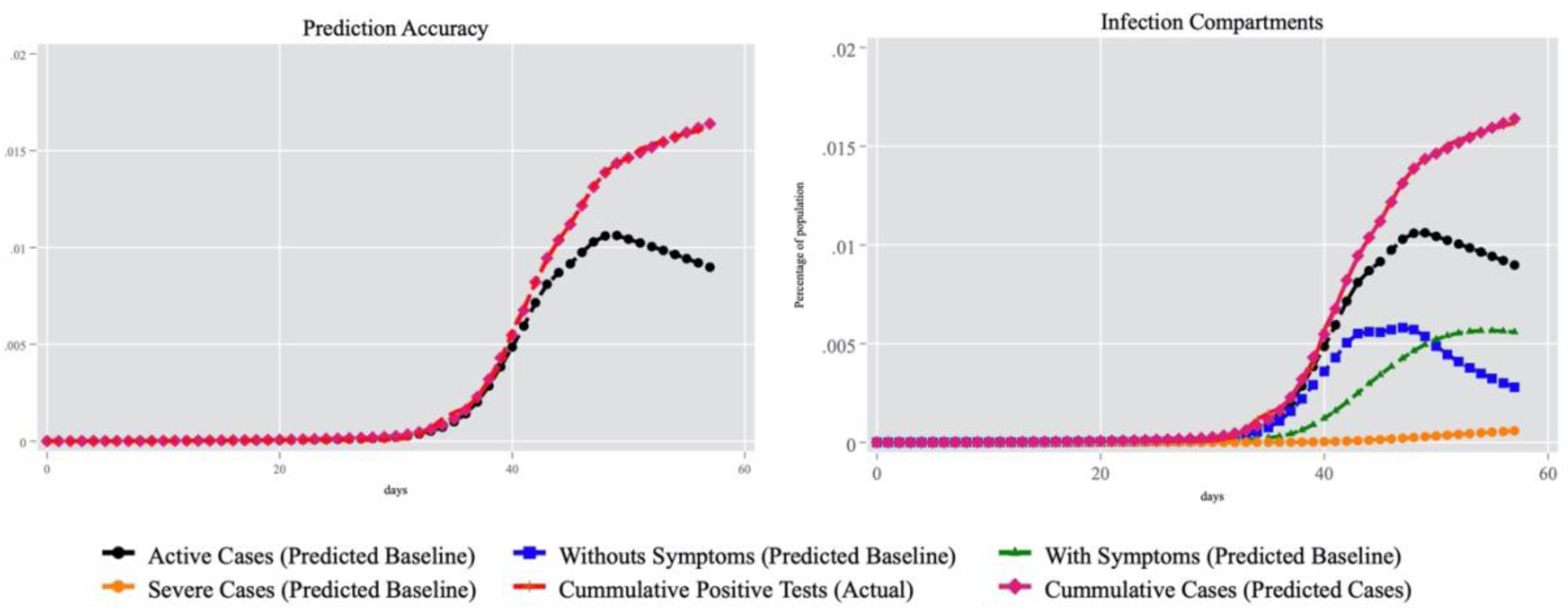
Benchmark, South Korea

For Ghana and Senegal, *R_t_* is tuned to match the number of official cases until the first reported case of community transmission (see table 1), that is the transmission that cannot be traced back to one of the initial cases. Then, *R_t_* is increased once and then lowered as soon as the first containment policy is enacted in the country and further lowered at each additional containment measure. Because Kenya first reported community transmission case coincided with the enforcement of a curfew to limit the spread of the virus, we only change *R_t_* once for both the community transmission and the curfew.

Since they alter *R_t_*, our baseline projections account for mitigation policies that were put in place in each of these countries (see figure 2). For instance, on the eighth day of the epidemic, Ghanaian officials restricted internal travels between infectious hot-spots and the rest of the country. Because these restrictions were announced 48 hours before they were effective, there is anecdotal evidence that a lot of individuals who lived in these hotspots travelled to other areas of the countries; we thus increase *R_t_* for two days, before decreasing it again when the internal limitations of travel were effective. Similarly, *R_t_* is tailored to each of the three countries according to the different policies they enforced. For example, in Kenya, we decrease it less for school closings than for regional lockdowns.

At the date of each containment measure, we adjust the value of *R_t_* and provide low policy and high policy effectiveness scenarios. Our baseline projections assume a moderate impact of the policy, while the high effectiveness projections correspond to the case in which containment measures reduce the reproduction number significantly — that is the situation in which the policy has had large positive impacts to reduce the effective rate of reproduction (see table 1). Our low policy effectiveness scenario translates the instance in which, on the contrary, the impact of each policy on the reproduction number is minimal.

**Figure 2:**
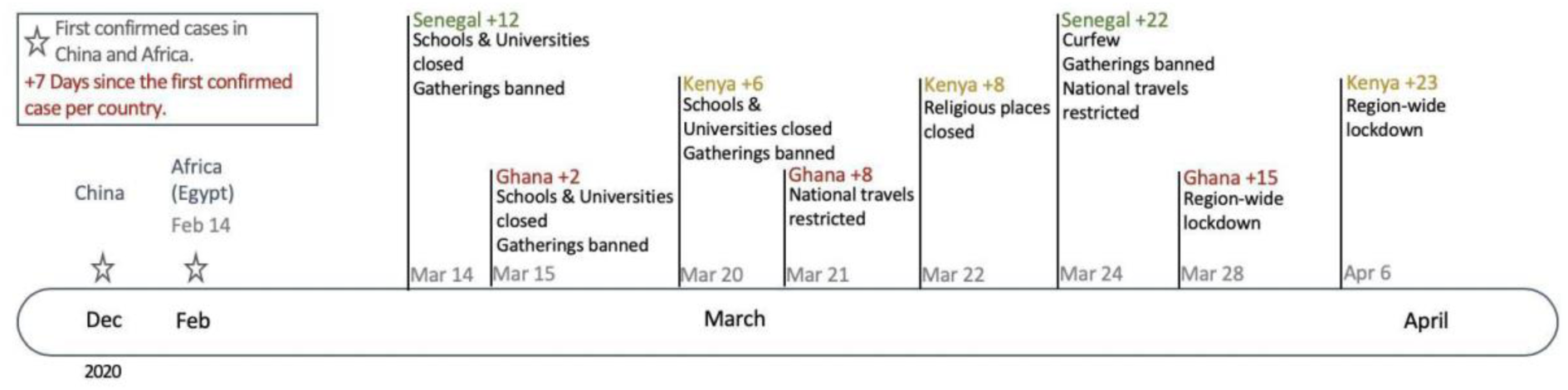
Timing of Policies Across Countries

**Table 1:** Parameters of the Model

|  | Ghana | Kenya | Senegal | South Korea | Source |
| --- | --- | --- | --- | --- | --- |
| 1st Case | 13-mars-20 | 14-mars-20 | 02-mars-20 | 20-janv-20 | European CDC (2020),<br>Sen. Ministry of Health (2020) |
| Pop. | 30.497 M | 51.808 M | 17.354 M | 51.410 M | CountryMeters[43] |
| $\mu_{birth}$ | 9.01E-05 | 9.71E-05 | 1.00E-04 | 2.51E-05 | CountryMeters[43] |
| $\mu_{death}$ | 9.01E-05 | 9.71E-05 | 1.00E-04 | 1.60E-05 | The World Bank [35] |
| $r_{as}$ | 0.4 | 0.4 | 0.4 | 0.4 | Nishiura et al. [17] |
| $T_{rec,as}$ | 14d | 14d | 14d | 14d | Hubei; McIntosh et al.[44] |
| $T_{inc}$ | 5d | 5d | 5d | 5d | McIntosh et al.[44] |
| $T_d$ | 6d | 6d | 6d | 6d | McIntosh et al.[44] |
| $T_{rec,ms}$ | 16d | 16d | 16d | 16d | McIntosh et al.[44] |
| $T_{sev}$ | 5d | 5d | 5d | 5d | McIntosh et al.[44] |
| $T_{rec,ss}$ | 18d | 18d | 18d | 18d | McIntosh et al.[44] |
| $\alpha$ | 2 | 2 | 2 | 2 | McIntosh et al.[44] |
| $d_{ms}$ | 0.37% | 0.37% | 0.37% | 0.03% | Country Meters[43] |
| $d_{ss}$ | 2% | 2% | 2% | 1.04% | Country Meters[43] |
| $r_{ms}$ | 98.34% | 98.53% | 98.62% | 94.33% | Verity et al. [45] |
| $R_t$ | 1.25, (1.0–1.4) if $0 \leq d < 7$<br>2.35, (2.2–2.7) if $7 \leq d < 9$<br>2.3, (1.7–2.5) if $9 \leq d < 16$<br>1.6, (1.0–2.2) if $16 \leq d$ | 2.4, (2.2–2.6) if $0 \leq d < 7$<br>2.15, (1.8–2.5) if $7 \leq d < 9$<br>2, (1.6–2.4) if $9 \leq d < 14$<br>1.6, (1.0–2.2) if $14 \leq d$ | 1.29, (1.1–1.48) if $0 \leq d < 20$<br>2.4, (2.1–2.7) if $20 \leq d < 26$<br>1.5, (1.0–2.00) if $26 \leq d$ | 1.13 if $0 \leq d < 30$<br>3.02 if $30 \leq d < 41$<br>1.85 if $41 \leq d < 44$<br>0.88 if $44 \leq d < 49$<br>0.3 if $49 \leq d < 59$ | Author's estimates, adjusting $R_t$ with timeline of govt. policies outlined in figure 2 |
| $\beta_{ms}$ | $0.41\beta_{as}$ | $0.41\beta_{as}$ | $0.41\beta_{as}$ | $0.41\beta_{as}$ | Author's estimate using Wang et al.[23] |
| $\beta_{ss}$ | $\beta_{as}/T_{rec,ss}$ | $\beta_{as}/T_{rec,ss}$ | $\beta_{as}/T_{rec,ss}$ | $\beta_{as}/T_{rec,ss}$ | |

Notes: $R_t$ in parenthesis are $R_t$ values for optimistic and pessimistic scenarios respectively.

Results reported in figure 3 show how the predictions fit the detected cases in the early days of the pandemic.

**Figure 3:**
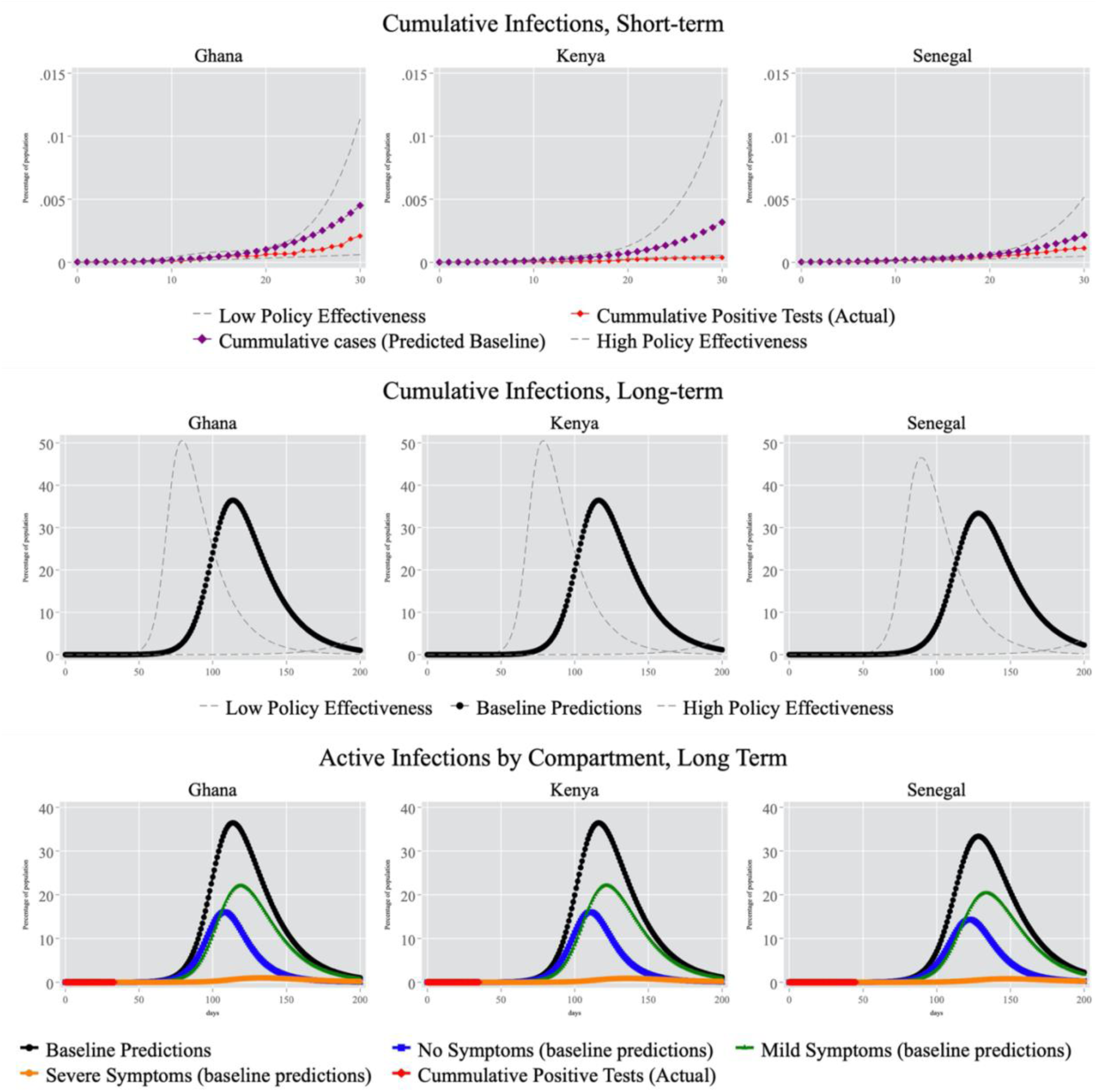
Projection of Active Infections

We report predictions for a year (see figure 3). Under the assumptions of the baseline model and their limitations, we predict that the peak of the epidemic will occur in July for all three countries as detailed in table 2. For Ghana, Kenya, and Senegal respectively, this peak should lead to approximately 11.1, 18.9, and 5.8 million active infections (including asymptomatic, and symptomatic cases) at the peak of the epidemic, with 308, 465, and 138 thousand individuals severely ill needing medical attention (see figure 3).

These long-term scenarios should be interpreted with great caution as they do not consider future policies or actions that could drastically reduce the contact rates and subsequently, flatten the curve further.^4^

**Table 2:** Projections of Active Cases at the Peak of the Epidemic for each Infected Compartment

|  |  | Active Cases at Peak |  |  |  |
| --- | --- | --- | --- | --- | --- |
|  |  | Days since first case | Severe symptoms | Mild symptoms | No symptoms |
| Ghana | Low Policy Effectiveness | 79 | 0.4M | 9.1M | 7.7M |
|  | Baseline | 114 | 0.3M | 6.7M | 4.9M |
|  | High Policy Effectiveness | 250 | 0.1M | 2.7M | 1.6M |
| Kenya | Low Policy Effectiveness | 79 | 0.6M | 15.4M | 13.1M |
|  | Baseline | 116 | 0.5M | 11.7M | 8.3M |
|  | High Policy Effectiveness | 252 | 0.2M | 4.6M | 2.7M |
| Senegal | Low Policy Effectiveness | 90 | 0.2M | 4.8M | 3.9M |
|  | Baseline | 128 | 0.1M | 3.5M | 2.5M |
|  | High Policy Effectiveness | 254 | 0.1M | 1.5M | 1.0M |

Note : The peaks of each infection sub-compartment might not align.

### Testing the Sensitivity of the Simulations to *R_t_*

We perform a sensitivity analysis for *R_t_* on our baseline model. We perturb it 100 times by *ε* drawn uniformly in [−5%*R_t_*, *+*5%*R_t_*]. The number of infections at the peak fluctuates between 27% to 40% of the total population in the active infection compartment for Ghana, between 28% and 40% for Kenya and between 25% to 37% for Senegal (see figure 2 of the appendix). In countries that enforced strict social distancing measures, predictions were significantly updated down — from about 2.2 million deaths on March 16,[12] to about 60 thousand on March 30.[26] A similar update can be expected from the outputs of our model as authorities take effective measures to reduce *R_t_* and/or people in these countries gradually adopt behavior that would minimize contacts.

### Population Density and Rate of Reproduction

As the population density increases, the rate of transmission of infectious diseases increases.[27] With respectively 43.3%, 72.2%, and 50.6% as a share of their population living in rural areas, Ghana, Kenya, and Senegal have sparsely populated areas outside of their main metropolitan areas, compared to countries like South Korea (18.5% of rural population).

There is little information on the relative rate of transmission of COVID-19 between rural and urban areas but we draw on other diseases, for which there is available data. During the 2014 Ebola outbreak in Sierra-Leone,[28] we find that the basic reproduction number in Kambia (the least densely populated district of Sierra-Leone) is .56 times the one of the Western Area Urban district (the most densely populated district, that comprises the capital Freetown)^5^. We take that to mean that the *R_t_* in rural areas was .56 times that of urban areas. Other mostly rural districts had a higher *R_t_*. To mirror the range of ratios of reproduction rates observed in mostly rural and mostly urban districts for the Ebola epidemic in Sierra Leone, we run two simulations, one in which the rural to urban ratio is .50, and one where it is .75. These two ratios bound the difference between mostly rural districts and mostly urban areas for Ebola in Sierra Leone.[28] Using these ratios, we calibrate the rural and urban reproduction rates so their population-weighted average is equal to the national *R_t_* which we keep constant across our baseline scenario and this scenario:

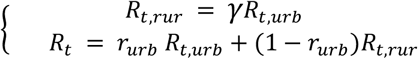

Where *R_t_,_rur_* and *R_t_,_urb_* are the rural and urban reproduction rates respectively; *γ=* .50, or *γ*=.75; *r_urb_* is the national urbanization rate; *R_t_* is the national reproduction rate listed in table 1. We use the first day of the first community transmission as the day of the first case in the rural area. Results are compiled in table 3. Effectively, we see in figure 4 that when accounting for rural areas, we observe two peaks. The first peak is driven by the spread in urban areas while the second peak, delayed in time is driven by the spread in rural areas. Kenya, with a rural share of the population of over 70% has the most noticeable split across its rural and urban areas.

### Co-Morbidity and Rise in the Occurrence of Severe Symptoms

Co-morbidity can impact the share of mild cases that develop severe symptoms.[29] In Asia and Europe, hypertension, obesity, diabetes, and coronary heart diseases have been drivers of adverse health outcomes.[29,30] Because the combined prevalence of diabetes, hypertension and obesity are not higher in Ghana, Kenya, and Senegal than they are in regions we use to derive the recovery rates, the baseline simulations already account for them. However, Ghana, Kenya, and Senegal have a persistent and high rates of anemia and tuberculosis[3] (see table 6). To our knowledge, there is no study on the magnitude of the impacts of anemia, tuberculosis, or HIV on the recovery of patients who have contracted the virus. We simulate two scenarios, with 25% and 75% of the recovery rate of otherwise healthy individuals for individuals with one of these underlying conditions1 (see table 5). In comparison, Zhou et al.[29] find that in Wuhan, China, patients with comorbidities (hypertension, diabetes, coronary heart disease, chronic obstructive lung disease, carcinoma, chronic kidney diseases and others) have a recovery rate equal to 73.2% of their otherwise healthy counterparts. Though uncertain for HIV, anemia and TB, the impact of these underlying conditions on the recovery of individuals will likely lie between these two bounds. This translates into adjusting *r_ms_* for individuals with TB, HIV, and anemia. Comorbidities have age-specific incidence rates; anemia affects women of child bearing age primarily, while HIV affects young adults at higher rates. The prevalence of HIV, TB and anemia are extracted from the open database Global Burden Disease.[31] We account for these age-based differences to compute the recovery rate of the population accounting for comorbidities:

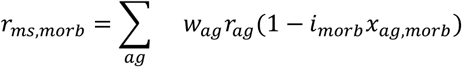

Where *r_ms,morb_* is the rate of recovery for infected individuals who develop mild symptoms and have one of the three comorbidities. *r_ag_* is the recovery rate of the otherwise healthy individuals in the age-group, *i_morb_* is either 1-.25 or 1-.75 depending on the scenario, and *x_ag,morb_* is the share of individuals in each age-group with the comorbidity.

We report the results in figure 4 and table 3. As expected, the predictions are higher in the case where we assume that individuals with comorbidities have a rate of recovery that is 25% that of otherwise healthy individuals. In the scenario with *R_t,rur_* = .75*R_t_,_urb_*, the number of active severe cases at the peak is 0.242M with a 75% recovery scenario for Ghana (0.308M for the 25% scenario), 0.313M for Kenya (0.631M) against and 0.104M (0.145M) for Senegal. Kenya’s large impact is driven by its larger HIV positive population.

**Table 3:** Projection of Infections Accounting for Africa Specific Factors

|  |  | Co-morbidity<br>% of the survival rate of healthy patients |  |  |  |  |  |  |  |
| --- | --- | --- | --- | --- | --- | --- | --- | --- | --- |
|  |  | 75% |  |  |  | 25% |  |  |  |
| | $\frac{R_t(rural)}{R_t(urban)}$ | Severe | Mild | Asympt. | Days* | Severe | Mild | Asympt. | Days* |
| Ghana | 50% | 0.2M | 4.4M | 3.3M | 81 | 0.3M | 4.8M | 3.9M | 81 |
|  | 75% | 0.2M | 4.8M | 3.9M | 96 | 0.3M | 4.4M | 3.3M | 96 |
| Kenya | 50% | 0.4M | 8.2M | 5.3M | 167 | 0.4M | 5.5M | 4.2M | 168 |
|  | 75% | 0.3M | 5.6M | 4.2M | 130 | 0.6M | 8M | 5.3M | 131 |
| Senegal | 50% | 0.1M | 2.2M | 1.6M | 87 | 0.1M | 2.3M | 1.9M | 87 |
|  | 75% | 0.1M | 2.4M | 1.9M | 105 | 0.1M | 2.1M | 1.6M | 105 |

\*Days of total infection peak, since the first case tested positive.
Note : The peaks of each infection sub-compartment might not align.

**Figure 4:**
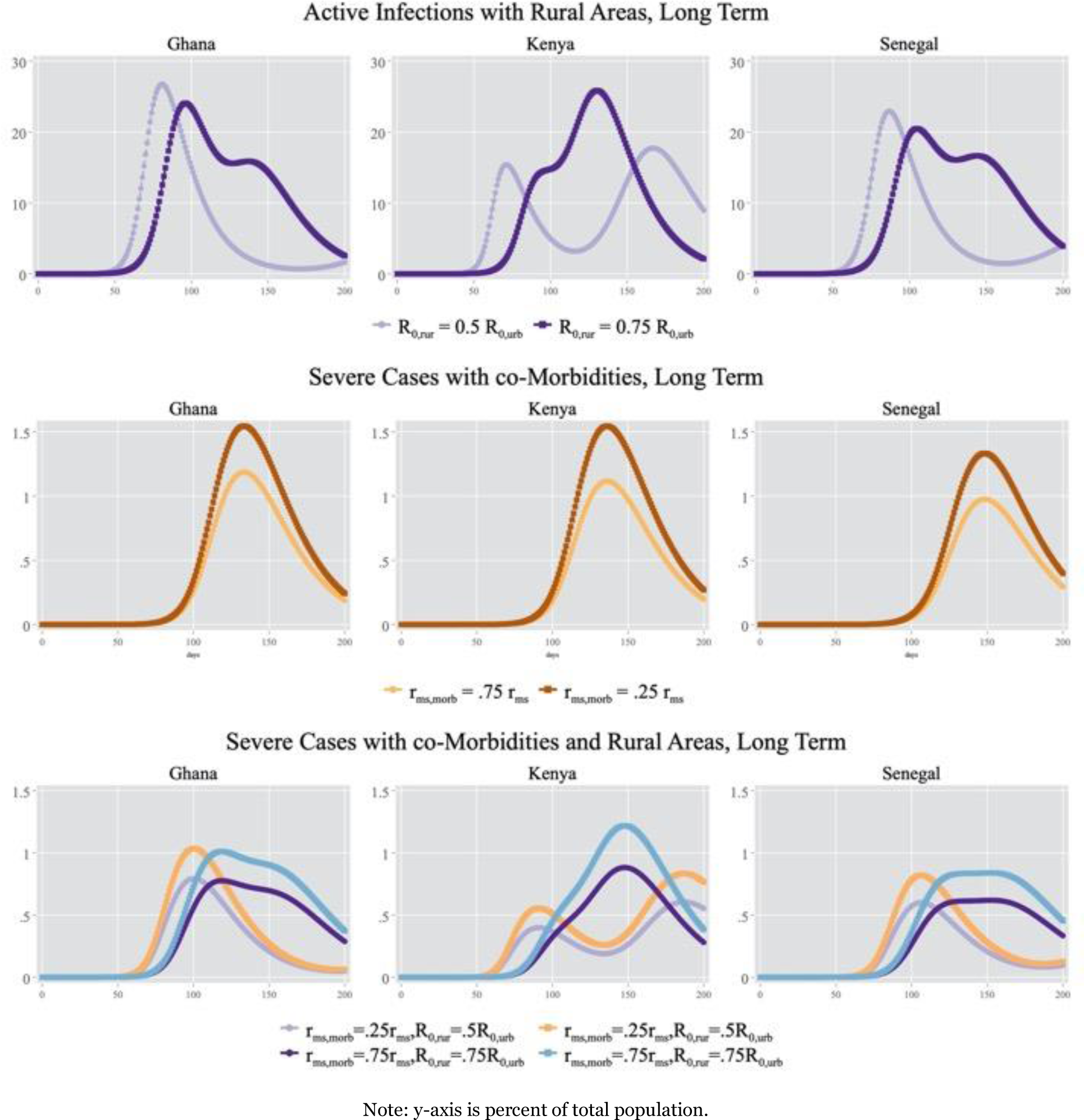
Projected Active Infections Accounting for Underlying Conditions and Rural Areas

### Mirroring South Korea’s effectiveness

Unlike countries in Europe, Ghana, Kenya, and Senegal have taken containment measures very early in the progression of the disease. The policies could have had impacts similar to the ones in South Korea. We present results of simulations mirroring the *R_t_* for South Korea. Specifically, we decrease *R_t_* for each country three weeks after the last recorded policy to 0.88, and then again at 6 weeks to 0.3. We find that the peak is much lower, with a number of active severe infections at the peak between 166 and 214 individuals for Ghana, 208 and 286 individuals for Kenya, and 140 and 189 individuals for Senegal; with the two bounds being a for recovery rates of respectively 75% and 25% of the recovery rate otherwise healthy patients. These peaks will occur two to three months after the first case (see figure 5). This scenario is attainable only if these countries are able to maintain effective policies for an extended period.

**Table 4:** Projection of Active Cases at Peak Accounting of Co-morbidities, With South-Korea’s *Rt*

|  |  | Co-morbidity<br>% of the survival rate of healthy patients |  |  |  |  |  |  |  |
| --- | --- | --- | --- | --- | --- | --- | --- | --- | --- |
|  |  | 75% |  |  |  | 25% |  |  |  |
| | $\frac{R_t(rural)}{R_t(urban)}$ | Severe | Mild | Asympt. | Days* | Severe | Mild | Asympt. | Days* |
| Ghana | 100% | 166 | 3,164 | 2,428 | 57 | 214 | 3,110 | 3,110 | 57 |
| Kenya | 100% | 208 | 4,221 | 3,206 | 60 | 286 | 4,134 | 4,134 | 60 |
| Senegal | 100% | 140 | 3,253 | 2,661 | 69 | 189 | 3,183 | 3,183 | 69 |

Note: The peak of each active cases for each sub -compartment might not align.

**Figure 5:**
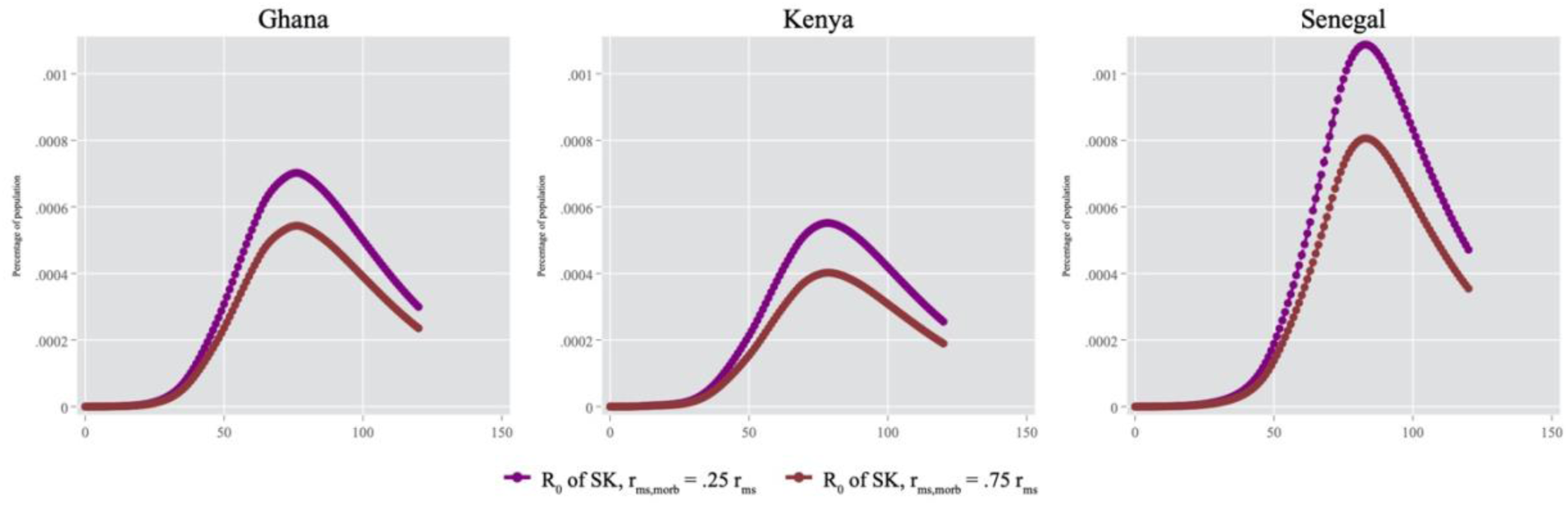
Projected Severe Active Infections Mirroring South Korean’s *R_t_*

## DISCUSSION

In this study, we account for the age structure of the population in each country, the burden of potential comorbidities and the differential spreads of the virus in rural and urban areas. We find that the relatively young population may limit the severity of the epidemic, by lowering the number of infections that lead to severe symptoms. We also find that sparsely populated areas may limit the spread of the epidemic. Rural areas effectively may lead to staggered peaks; this has important implications for policy makers who may be faced with two waves, and so may need to adapt their responses to adaptively deploy personnel on their territory as these peaks occur.

High rates of comorbidities however may lead to more individuals developing severe symptoms relative to a scenario with no comorbidities. We find that at the peak Ghana, Kenya, and Senegal are predicted to have respectively between 0.78 and 1.03%, 0.89 and 1.22%, and finally, 0.60 and .84% active clinical severe cases of COVID-19 with a peak of total infections predicted to occur between June 2 and June 17 (Ghana), July 22 and August 29 (Kenya), and May 28 and June 15 (Senegal) respectively against a July timeline for our baseline specification. Successful containment policies could lead to even lower rates of severe infections.

Though recent models look at a few countries in Africa,[32] or at the continent as a whole,[33,34], there are little to no studies predicting the spread of COVID-19 in Ghana and Senegal while incorporating specificities of these two countries. In Kenya, however Brand et al.[10] account for age-based population mixing and assume that asymptomatic individuals are as infectious as symptomatic individuals to predict that by the end of the year, 46.1 million (i.e. 89% of the public) of infections will have occurred. This prediction is comparable with the baseline results of our study in the absence of further containment policies. In that scenario, we find that about 47.7 million (93% of the public) individuals may be infected cumulatively.

Containment measures will be successful only if the public complies; however, measuring compliance is complex and has not been rigorously studied in the context of COVD-19 in these countries. In Ghana, Kenya and Senegal poverty is the main challenge to compliance, with official unemployment rates reaching 68.7%, 51.3% and 64.6% respectively.[35] As a response, authorities have implemented emergency transfer programs in cash and in-kind to the most vulnerable households partly to address compliance but also to avoid a humanitarian crisis (Senegal, Ghana). In urban areas, officials have required buses and taxis to reduce their number of passengers (Kenya, Senegal) and have mandated the use of masks (Ghana, Senegal).

Looking at how spike in cases was met by various healthcare systems in Europe and Asia, it is likely that most asymptomatic and mild cases may remain undetected.

### Limitations

Our model does not incorporate changes in the survival rate of the virus due to weather or humidity, and in that regard, our simulations are a worst-case scenario.[36,37] Additionally, the model assumes homogeneous mixing of individuals within rural areas and urban areas which is an unlikely assumption. In a future iteration of our model, we plan to use a spatially-structured model in order to relax the homogeneous mixing assumption by leveraging phone data.[38–40]

The model also excludes international population flows. All countries in our sample have closed their international borders — airports and roads — before or a few days after their first confirmed imported case (see figure 2). However, it is possible that COVID-19 was spreading undetected for days in the respective countries. If that is true, the peak of active cases might be delayed in comparison to the true peak. Furthermore, the spread of this disease is highly dependent on the reproduction number *R_t_*. Since this number is contingent upon many factors (policies, individuals’ behavior etc.); its value in the long-run is subject to large uncertainties. The projected number of infections in the medium to long term could thus be considerable overestimates (or underestimates) of the true number of infections (depending on the scenarios).

These predictions aggregate infections in rural and urban areas, however, in practice, the peaks in urban areas, due to higher reproduction rates, will occur earlier. In rural areas however, the peaks will be delayed due to their lower *R_t_*. This distinction is important for policy makers who can target their resources accordingly.

The use of the data also comes with limitations such as the inaccuracy of the data collection. For example, one person was tested positive for COVID-19 on March 4 but entered Senegal on February 24. We expect that all the countries in our sample are dealing with similar delays, however, we did not find a consistent way to address this issue. Additionally, given the low number of tests performed to detect the virus, we cannot ex-ante measure the accuracy of our model in Ghana, Kenya, and Senegal.

Because outcomes of individuals with critical needs are highly dependent on the capacity of health care systems, having data on health care capacity is important in predicting the number of fatalities. In our simulations, information such as the number of intensive care unit beds would inform the fatality rate of individuals with severe symptoms (*d_ss_*). Unfortunately, we do not have access to such data and we thus choose not to show the results for fatalities rates in these three countries. Finally, we use a SIR, which assumes perpetual immunity – however, there are still uncertainties regarding the possibility of reinfection.[41]

## CONCLUSION

In conclusion, containment measures, age structures, low urbanization and co-morbidity may lead Ghana, Kenya, and Senegal to having different trajectories from the USA, and from Asian and European countries. This study is a first attempt at accounting for rural densities and co-morbidity, and it suggests that rural areas will slow down the spread of the epidemic, and that relatively young population will keep the number of severe cases low compared to the nearly 3.5% hospitalization rate in Europe and central Asia.[42] Our findings also show how sensitive these results are to different assumptions on the effectiveness of policies, assumptions on co-morbidities and differential effective rates of reproduction in rural and urban areas.

## Data Availability

All data used for the purpose of this study are aggregated and publicly available.

## Appendix

**Figure 1:**
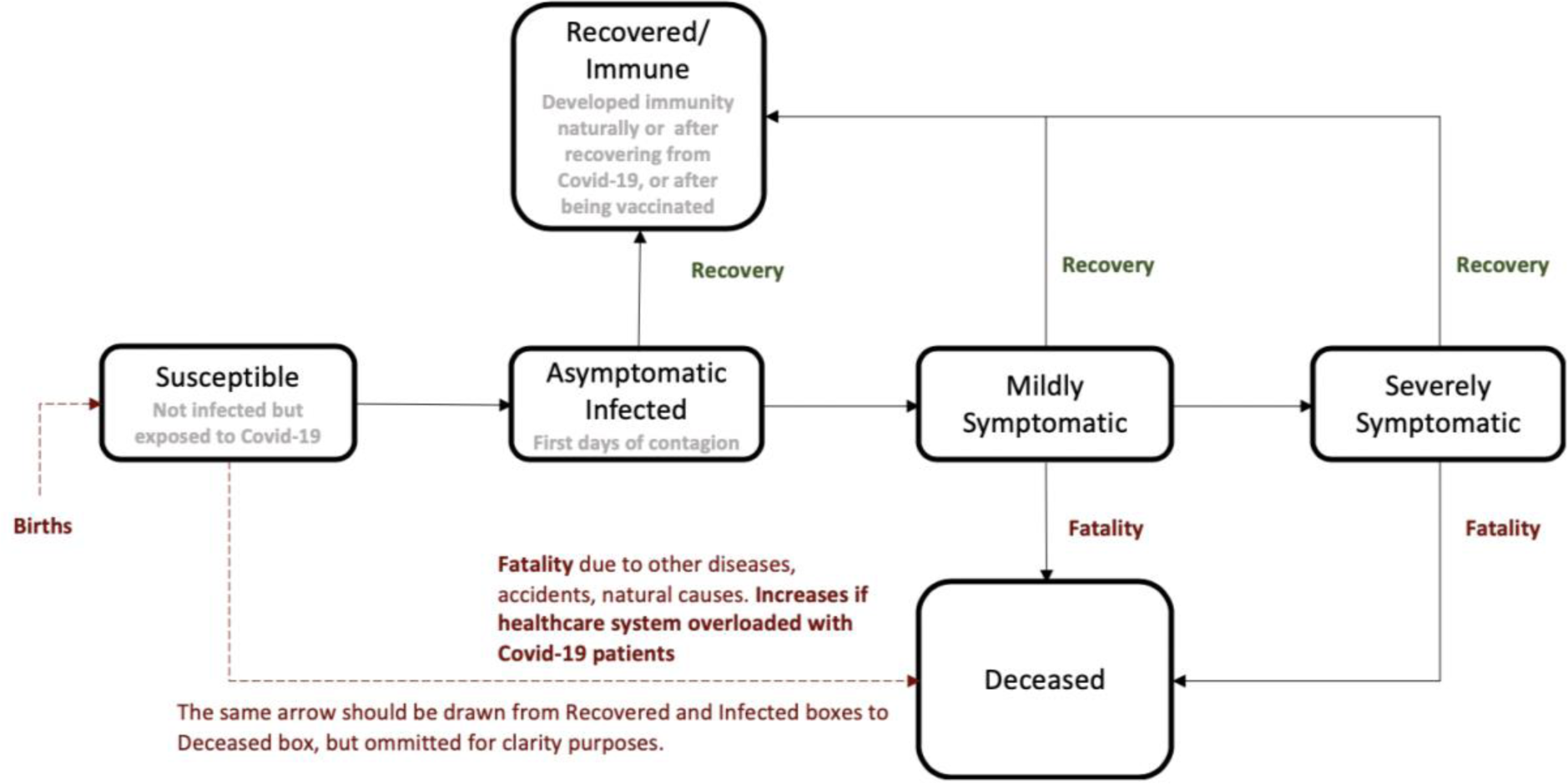
Visual representation of the model

**Figure 2:**
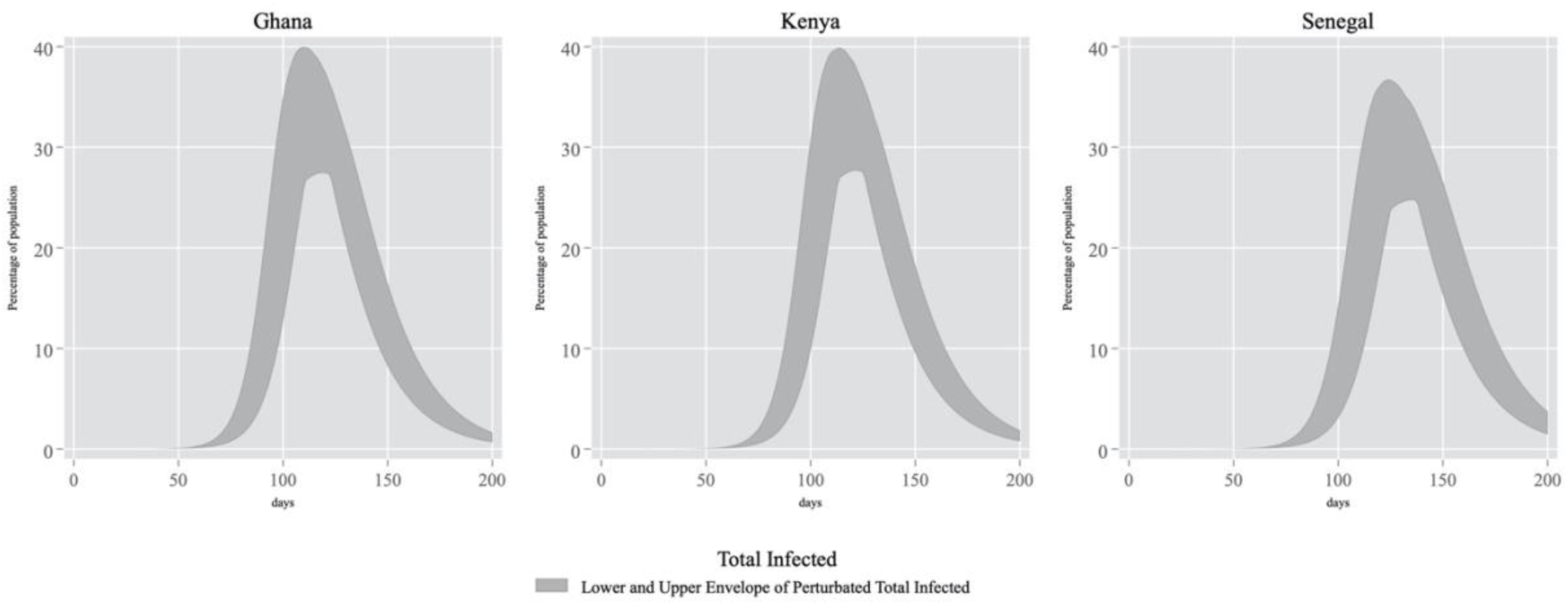
Sensitivity of Active Cases to Perturbations of *R_t_*

**Table 1:** Projection of Active Cases Accounting for Co-morbidity or Rural/Urban factors

|  |  |  | Active cases at peak |  |  |  |
| --- | --- | --- | --- | --- | --- | --- |
|  |  |  | Severe | Mild | Asympt. | Days* |
| Ghana | Rural/Urban** | 50% | 0.2M | 4.8M | 3.9M | 81 |
|  |  | 75% | 0.2M | 4.4M | 3.4M | 96 |
|  | Co-morbidity*** | 25% | 0.5M | 6.6M | 4.9M | 114 |
|  |  | 75% | 0.4M | 6.7M | 4.9M | 114 |
| Kenya | Rural/Urban | 50% | 0.2M | 5.7M | 4.2M | 167 |
|  |  | 75% | 0.4M | 8.2M | 5.3M | 130 |
|  | Co-morbidity | 25% | 0.8M | 11.3M | 8.3M | 116 |
|  |  | 75% | 0.6M | 11.4M | 8.3M | 116 |
| Senegal | Rural/Urban | 50% | 0.08M | 2.4M | 1.9M | 87 |
|  |  | 75% | 0.09M | 2.2M | 1.6M | 105 |
|  | Co-morbidity | 25% | 0.2M | 3.5M | 2.5M | 129 |
|  |  | 75% | 0.2M | 3.5M | 2.5M | 128 |

\* Days of total infection peak, since the first case tested positive.
\*\* $R_t(rural)$
$R_t(urban)$
\*\*\*Recovery rate for patients with preexisting conditions as percentage of healthy patients' recovery rates.
Note: The peak of each active cases for each sub -compartment might not align.

## Footnotes

1 As of May 1st, 2020, Ghana has conducted 3.37 tests per thousand individuals while Kenya and Senegal are respectively at 0.4 and 0.76.[11]

2 As of April 25, 2020, the Western African fatality rate is 2.49% while the Eastern African fatality rate is 2.25%, we take the average of both these numbers. Note that we use the regional figures because the number of cases at the national level is still low in the three countries. Western Africa has 8,034 cases and Eastern Africa has 3,319 cases as of April 27, 2020 (Africa CDC). Although these numbers are much lower than the Africa-wide (32,182 cases) and the worldwide ones, we find them more appropriate as they are more faithful to the standards of living and the age pyramid of the countries we study. Particularly, Algeria (12.57% fatality rate) and Egypt (6.99%), among others, raise the Africa-wide fatality rate to 4.44% but are structurally different from Ghana, Kenya and Senegal. That being said, we acknowledge that our choice relies on the testing capacity in both Western and Eastern African regions and might underestimate the true fatality rate as a consequence.

3 Note that there are two ways to compute the fatality rate, either (1) as the ratio deaths / total cases, or (2) deaths / closed cases. While the former is likely to be an underestimate because lots of open cases can still end up in death, the latter is an overestimate because it’s likely that deaths are closed quicker than recoveries. As critical cases are more likely to be detected than mild infections, it is also likely that the number of true cases is underestimated by official numbers but the number of COVID-19 related deaths is relatively well captured by official reports. Therefore, the ratio (1) is likely to be a better estimate of the true fatality rate than (2), and we use this definition of fatality rate.

4 For instance, wearing a mask in public space is now mandatory in Kenya (since April, 15), Senegal (since April, 20) and Ghana (since April, 25). Additionally, the government of Ghana lifted its partial lockdown on April, 20.

5 We chose the most and the lease populated districts, because all these districts include urban areas.

